# Digital cytometry of immune cell types from dried blood microsamples

**DOI:** 10.64898/2026.11.13.26368027

**Authors:** Quentin R. V. Ferry, Timothy Rajakumar, Marco Zeeck, Vanessa Chong-Morrison, Tammo von Schrenck, Keihan Ahmadi-Simab, Bruno R. Steinkraus

**Affiliations:** Muno Inc., MA, USA; Muno GmbH, Hamburg, Germany; Medizinicum, Hamburg, Germany; Tidy Bio Ltd, Oxfordshire, United Kingdom; Labor Dr. von Froreich, Hamburg, Germany

**Keywords:** blood, microsampling, deconvolution, transcriptomics, cytometry

## Abstract

Enumeration of circulating immune cells is an established clinical practice implemented across a plethora of indications. While flow cytometry is the gold standard for this task, cellular deconvolution algorithms provide a robust computational alternative using molecular data. These digital cytometry pipelines typically rely on millilitre-scale venous blood samples. Here, we evaluated the potential of capillary blood microsampling as a patient-centric and minimally invasive approach for the digital cytometry of major lymphocyte populations (T cells, B cells, and natural killer cells; TBNK). In a cohort of 200 participants, we compared RNA sequencing based deconvolution performance between matched PAXgene-stabilised venous blood and Mitra capillary blood microsamples. While venous transcriptomes enabled highly accurate TBNK estimation, patient-matched capillary microsamples exhibited a significant reduction in deconvolution accuracy. Computational adaptations, including the integration of capillary-specific single-cell reference profiles, yielded only limited improvements. Instead, our analyses revealed that the performance deficit was primarily driven by the decreased transcriptome complexity of the capillary microsample libraries. This study highlights the critical role of transcript diversity for cellular deconvolution and reveals the current pre-analytical challenges that must be overcome to harness transcriptomics based digital cytometry using dried blood microsamples.

## 1. Introduction

The composition of circulating immune cells provides clinically valuable information for disease diagnosis, patient stratification, and longitudinal health monitoring. Flow cytometry is commonly used to measure immune cell populations directly but requires intact cells, specialised equipment, and time-sensitive sample processing. Digital cytometry provides a complementary computational approach in which corresponding cell type fractions are estimated from molecular measurements, including transcriptomic [1] and epigenetic data [2]. When combined with appropriate sample stabilisation, this approach is compatible with remote collection, storage, shipping, and delayed analysis. A wide range of bioinformatic methods have been developed to deconvolve bulk RNA sequencing (RNA-seq) and DNA methylation datasets into their constituent cell types.

Previous studies have demonstrated robust deconvolution of venous whole blood transcriptomes into their constituent immune cell populations, with strong correlations between computationally estimated cell fractions and flow cytometric measurements [3, 4, 5, 6, 7]. These studies have generally relied on millilitre-scale venous blood samples collected by trained personnel through venipuncture. Capillary blood microsampling, however, typically requires 120 µL or less and offers a minimally invasive alternative that is well suited to repeated, out-of-clinic, and home-based collection [8, 9].

Studies have shown that capillary microsamples can support reliable measurements of metabolites [10], proteins [11, 12], and drugs [13, 14] and can generate global transcriptomic profiles broadly comparable to those obtained from venous blood [15]. However, lower RNA input, differences in collection and processing, and biological differences between capillary and venous blood may affect the accuracy of transcriptome-based cell type deconvolution.

To evaluate the potential of capillary blood microsampling for digital cytometry in diagnostics and health monitoring, we investigated whether the major lymphocyte populations comprising T cells, B cells, and natural killer cells (TBNK) could be reliably deconvolved from capillary blood RNA-seq data. We recruited 200 participants and generated paired transcriptomic profiles from capillary microsamples and venous whole blood samples. RNA-seq-derived estimates of TBNK cell fractions were evaluated against matched flow cytometric measurements from the same participants. This paired design allowed us to assess the accuracy of capillary blood deconvolution and compare its performance with that achieved using conventional venous samples.

## 2. Materials and Methods

### Patient recruitment and ethics

The Muno-1 study was a single centre, descriptive study conducted at the rheumatology outpatient department of the Medizinicum Hamburg. Adults aged 18 years or older without known chronic infection with HIV, hepatitis B virus, hepatitis C virus, or anaemia, were eligible. Recruitment continued until 200 participants had been fully enrolled. Participants provided written informed consent before completing the study questionnaire and undergoing blood collection. Samples and associated clinical data were pseudonymised at the study site before transfer for laboratory analysis, with the reidentification key retained at the clinic. The study was conducted in accordance with the Declaration of Helsinki and the professional code of the Hamburg Medical Association and was approved by the Ethics Committee of the Hamburg Medical Association on 15 May 2024 (reference: 2024-101263-BO-ff).

### Study design

The study used a paired, within-participant design. At a single study visit, each participant underwent venous blood collection and finger-prick capillary blood microsampling. Three matched sample modalities were generated: fresh venous whole blood for differential blood counting and flow cytometric measurement of TBNK populations; PAXgene Blood RNA Tube (BD Biosciences, catalogue no. 762165)-stabilised venous whole blood (two tubes) for RNA-seq; and dried capillary blood collected using Mitra devices with VAMS technology (Trajan Scientific, VM-304KMBD) (four 30 µL tips) for RNA-seq. Fresh blood was collected in EDTA vacutainers and sent for same-day flow cytometry analysis at Labor Dr. von Froreich, Hamburg. PAXgene samples were immediately frozen at *−*20 ^°^C for short-term storage before transport and longer term storage at *−*80 ^°^C. Mitra samples were dried and resealed in their desiccant pouch according to the manufacturer’s instructions. The samples were stored at *−*20 ^°^C short-term followed by long-term storage at *−*80 ^°^C until processing.

### Flow cytometry

Fresh EDTA-anticoagulated venous whole blood was analysed on the day of collection at Labor Dr. von Froreich, Hamburg, using an automated AQUIOS flow-cytometry system (Beckman Coulter). Samples were mixed immediately before loading and were processed automatically by the instrument using the AQUIOS Tetra assay.

### Stabilised peripheral blood RNA extraction

One PAXgene Blood RNA Tube per patient was thawed overnight before total RNA extraction using QIAGEN Genomic Services’ standard 96-well plate-based extraction workflow for PAXgene-stabilised blood. RNA concentration was determined by optical-density measurement, and RNA integrity was assessed using an Agilent TapeStation and reported as the RNA integrity number equivalent (RINe).

### Dried capillary blood microsample RNA extraction

Total RNA was extracted from two 30 µL Mitra tips using the NucleoMag RNA kit (Macherey-Nagel, 744350.4) according to the manufacturer’s instructions. The tips were dispensed into Lysis Buffer and vortexed at maximum speed for 10 s at room temperature to release RNA. Lysates were cleared by centrifugation using Investigator Lyse & Spin Basket Kit (Qiagen, 19598) prior to further processing on a MagnetaPure 32+ automation system. RNA was eluted in 50 µL volume and quantified using Quant-iT RNA HS (ThermoFisher, Q32884). RINe was measured using Agilent TapeStation HS RNA (Agilent, 5067-5579).

### RNA-seq library preparation and sequencing

Library preparation was performed using QIAseq UPX 3’ Transcriptome Kit (QIAGEN, 333088). 10 ng (PAXgene) or 1–2 ng (Mitra) purified RNA was converted into cDNA NGS libraries. Library preparation was quality controlled using capillary electrophoresis with Agilent DNA 7500 (Agilent, 5067-1506) on a 2100 Bioanalyzer before the samples were pooled in equimolar ratios. The library pool(s) were quantified using qPCR and sequenced on a NextSeq 2000 (Illumina Inc.) sequencing instrument to an average read depth of 10 000 000 reads per sample. Raw data was demultiplexed and FASTQ files for each pool were generated using Illumina bcl2fastq version 2.20.0.422.

### Read mapping, demultiplexing, and count table generation

Raw FASTQ reads were mapped by STARSolo version 2.7.10b [16] using default parameters and a whitelist of QIAGEN barcodes for demultiplexing. QIAGEN barcodes were matched to their known patient IDs to generate the final count matrices.

### *In silico* enumeration of major lymphocyte cell types

#### Bulk RNA-seq cellular deconvolution

Count matrices were imported into R 4.5.1/RStudio 2024.12.1+563 running in an Ubuntu 22.04.5 LTS environment. Principal-component Analysis (PCA) identified 1 PAXgene outlier and 4 Mitra outliers. A further 26 Mitra samples with asymmetrical skew in the distribution of their normalised gene counts were also excluded. With the retained 199 PAXgene and 170 Mitra samples, gene counts were normalised using the trimmed mean of M-values (TMM) method to compute Counts Per Million (CPM) values as implemented in edgeR [17]. CIBERSORT [3], EPIC [4], and quanTIseq [5] deconvolution were performed using immunedeconv [18] R package distributed via Bioconductor version 3.19. Ground truth (i.e., “measured”) fractions of whole blood per cell type (T cells shown as an example) were computed from flow cytometry measurements as follows– *T*.*Cells*.*rel*(*relative*, %)/100 *× Lymphocytes*.*rel*(*relative*, %). Custom reference profiles for EPIC deconvolution were generated from capillary blood scRNA-seq datasets [19] using the Seurat version 5.2.1 ecosystem [20]. Highly variable features were identified using scTransform [21] implemented within Seurat.

#### Feature selection using genetic algorithm

To identify minimal gene sets capable of accurate TBNK deconvolution, a genetic algorithm-based feature selection approach was implemented. The algorithm operated on normalised gene expression matrices (log_2_(CPM + 1) transformation) from the PAXgene datasets to identify optimal gene subsets for predicting flow-cytometry-derived TBNK fractions.

The genetic algorithm maintained a population of 500 candidate gene sets (individuals), where each individual represented a unique combination of *n* = 24 genes. Initial populations were randomly seeded from the full gene expression matrix. Fitness evaluation for each gene set employed 50-fold cross-validation using linear regression models, with fitness scores calculated as the product of Pearson correlation coefficients across TBNK predictions (Fitness 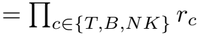 This multiplicative fitness metric ensured balanced performance across all three cell types.

Evolution proceeded through iterative generations combining selection, crossover, and mutation operations. In each generation, the 500 fittest individuals were retained based on their fitness scores. Breeding generated 500 offspring through weighted probabilistic selection of parent pairs (exponentially decaying probability: *p*_*i*_ ∝ *e*^*−*6*i*/*N*^ where *i* represents rank position), with each offspring inheriting genes randomly sampled from both parents. Mutation was applied to 20 % of genes in each offspring to maintain genetic diversity. Additionally, 250 randomly generated individuals were introduced each generation to prevent premature convergence.

To ensure robust feature selection, a leave-one-batch-out cross-validation scheme was employed. The dataset was partitioned into 20 equal batches, with each instance of the genetic algorithm tested on a specific batch after training on the union of the remaining 19. This process ran for 1000 generations per batch, with parallel processing across four cores to reduce computational time. For each run, we confirmed that the fitness level within the population had plateaued for at least 50 generations prior to termination. Final gene importance scores were calculated as the frequency of selection across all 20 independent runs, providing a consensus ranking of predictive genes for TBNK deconvolution. The algorithm implementation utilised scikit-learn [22] version 1.5.2 for machine learning components and multiprocessing for parallelisation.

## 3 Results

### Cohort characteristics

A total of 200 adults were enrolled in the Muno-1 study. Disease-stratified cohort characteristics were available for 192 participants, of whom 65 were classified as having no recorded disease and 127 as having one or more recorded diseases; eight participants were omitted from these stratified summaries because of missing information or an incomplete year of birth. Participants in the disease group were older than those in the no-disease group, with mean ages of 46.9 and 36.6 years, respectively, and had a higher mean body mass index of 26.1 kg m*−*2 compared with 23.1 kg m*−*2. Women comprised the majority of both groups. Active autoimmune disease was the most frequently recorded disease category, affecting 31.4 % of participants in the disease group. Full demographic, clinical, and lifestyle characteristics are presented in Table 1.

**Table 1:**
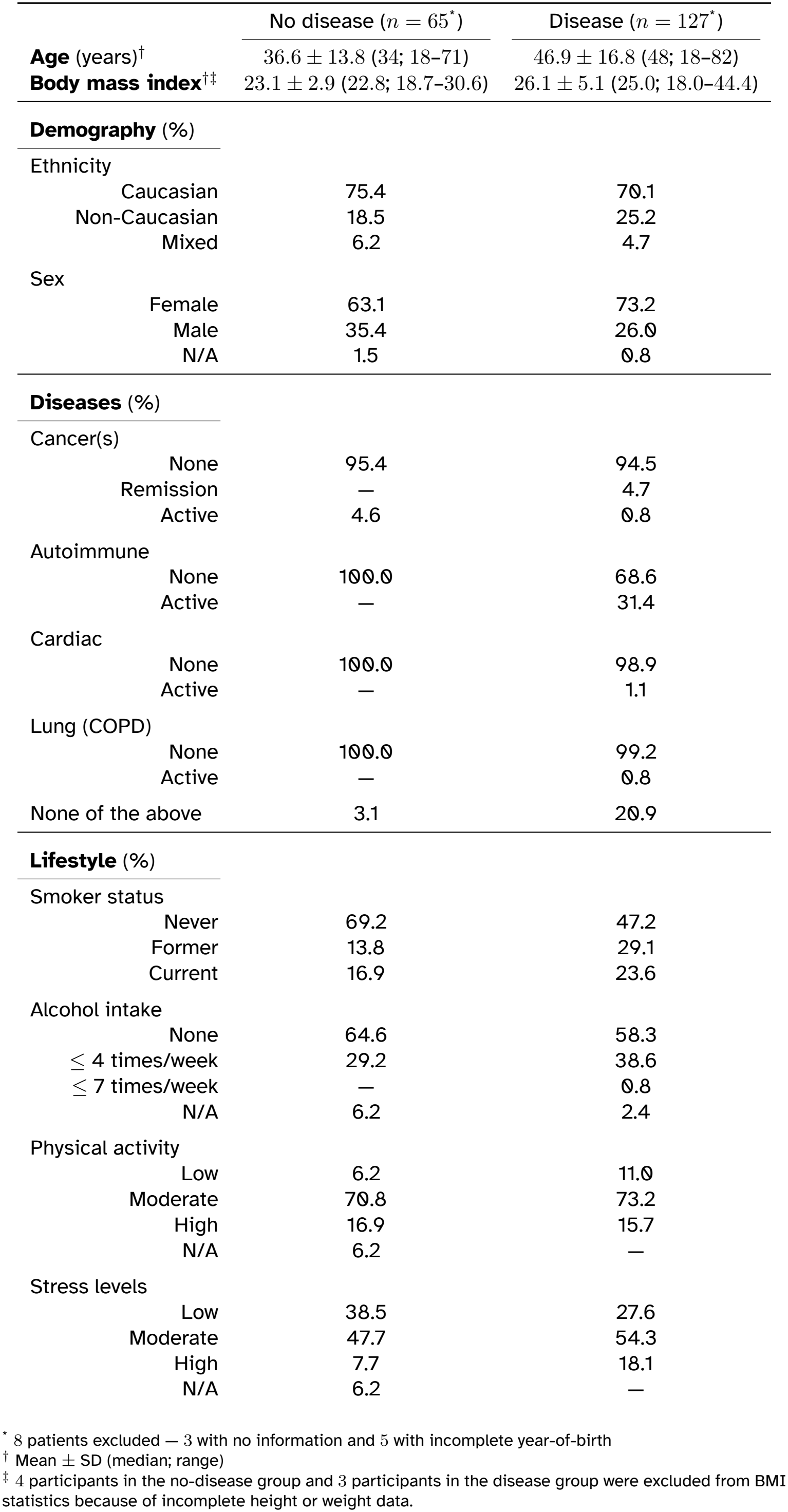
Muno-1 cohort characteristics (*N* = 200)

### Digital cytometry of TBNK fractions from stabilised venous blood

We first evaluated whether RNA-seq profiles generated from PAXgene-stabilised venous blood were compatible with established digital cytometry methods (Figure 1A). Principal-component analysis (PCA) identified one PAXgene library as an outlier, which was excluded, leaving 199 samples for analysis (Figure 1B). The retained libraries contained a median of 15 786 detected genes per sample. Prior to deconvolution, the expression matrix was restricted to 8844 annotated genes detected in all the retained 199 samples.

**Figure 1:**
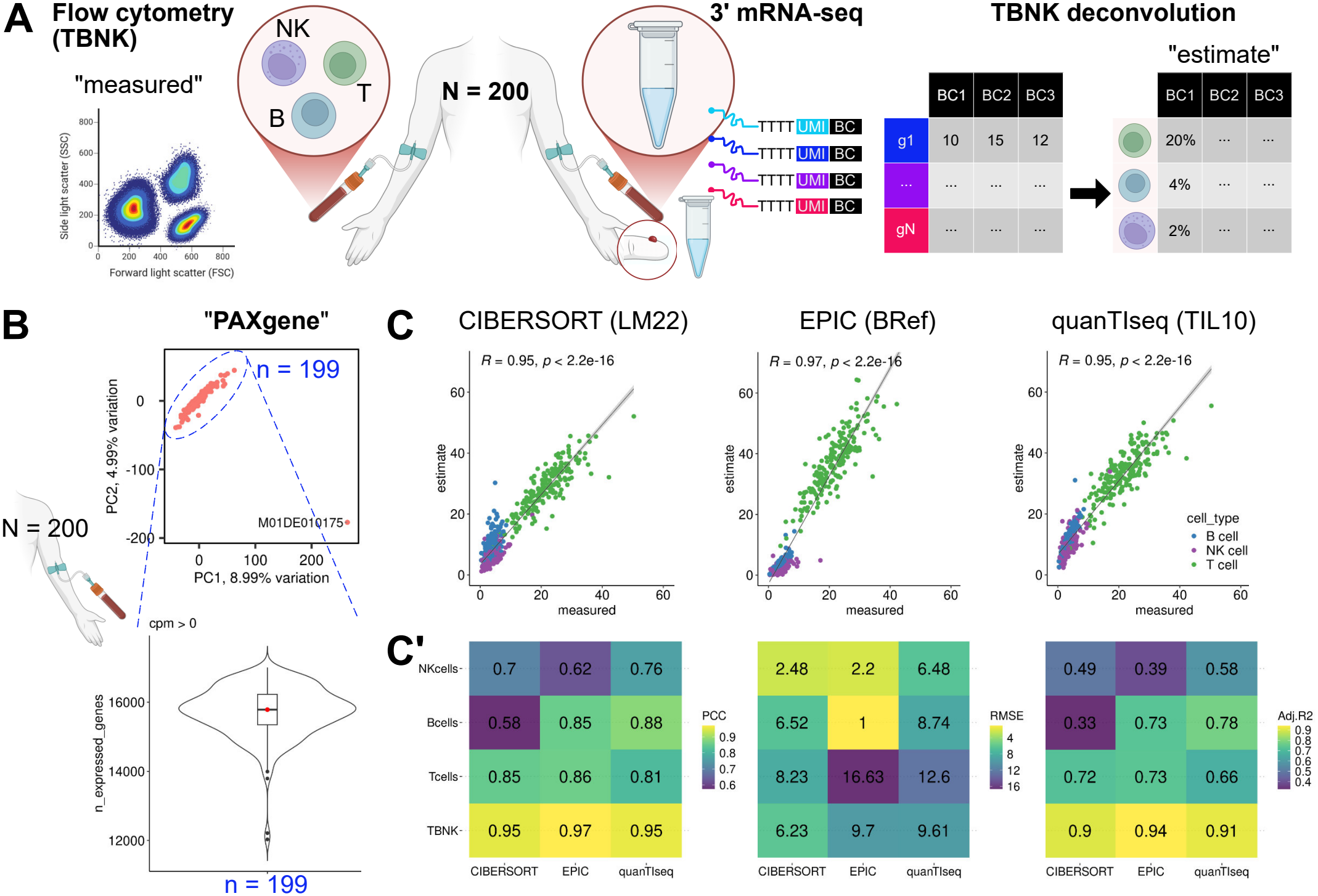
Study design and validation of RNA-seq workflow for TBNK deconvolution task. **(A)** Three blood draws per individual were collected from a cohort of 200 patients – venous fresh blood for flow cytometry, venous stabilised blood, and dried finger-prick capillary blood. Bulk 3’ RNA-seq was performed on venous stabilised blood and dried finger-prick samples to generate *gene*(*g*) *×patient*(*BC*) UMI-corrected count matrices. TBNK fractions were estimated from gene counts using cellular deconvolution. **(B)** PCA of venous stabilised blood (“**PAXgene**”) dataset with 1 outlier excluded. Violin plot of detected genes (CPM *>* 0) in remaining 199 samples, with median (15 786 genes) highlighted in red. **(C)** Scatter plots of computed TBNK fractions (“estimate”) vs ground-truth flow cytometry counts (“measured”) using three deconvolution algorithms (CIBERSORT, EPIC, quanTIseq) and their respective pre-packaged signatures (LM22, BRef, TIL10). Pearson’s *r* and *p* value for collective TBNK deconvolution results are labelled. **(C’)** Tile plots summarising performance metrics of TBNK deconvolution split by cell type (“Tcells”, “Bcells”, or “NKcells”) or collectively (“TBNK”), across the three algorithms. PCC; Pearson Correlation Coefficient, RMSE; Root Mean Squared Error, Adj.R2; Adjusted R-squared, NK; Natural Killer.

TBNK cell fractions were estimated using CIBERSORT [3], EPIC [4], and quanTIseq [5]. All three methods produced estimates that positively correlated with the matched flow cytometric measurements, although performance varied by cell type and method (Figure 1C–C’). As detailed in the performance heatmaps (Figure 1C’), quanTIseq showed the most consistent cell type specific performance across the Pearson correlation coefficient, adjusted R^2^, and root-mean-square error (PCC = 0.76–0.88; Adj. R^2^ = 0.58–0.78; RMSE ≤ 12.6 %). CIBERSORT performed least well for B cell fractions (PCC = 0.58; Adj. R^2^ = 0.33), whereas EPIC performed least well for NK cell fractions (PCC = 0.62; Adj. R^2^ = 0.39). Although EPIC achieved a strong correlation for T cell fractions, it showed comparatively high prediction error (RMSE = 16.6 %). When observations from the three cell types were considered collectively, all methods achieved strong overall correlations and low prediction error (PCC ≥ 0.95; RMSE ≤ 9.7 %). These results established that the PAXgene RNA-seq dataset was suitable for subsequent comparison with dried capillary blood microsamples.

### Digital cytometry accuracy is reduced in dried capillary blood microsamples

We next evaluated deconvolution performance using the participant-matched Mitra dried capillary blood microsample transcriptomes. PCA identified 4 outlier libraries, and a further 26 libraries showed abnormal distributions of normalised gene counts (Figure 2A). Exclusion of these 30 libraries resulted in a quality-controlled analysis set of 170 samples. The retained Mitra libraries contained a median of 9600 detected genes per sample (Figure 2A, right), substantially fewer than the median of 15 786 genes detected in the PAXgene libraries. To account for the observed variability in gene dropout between Mitra samples, prior to deconvolution, the expression matrix was restricted to 8882 annotated genes detected in at least 50 % of the retained 170 samples.

**Figure 2:**
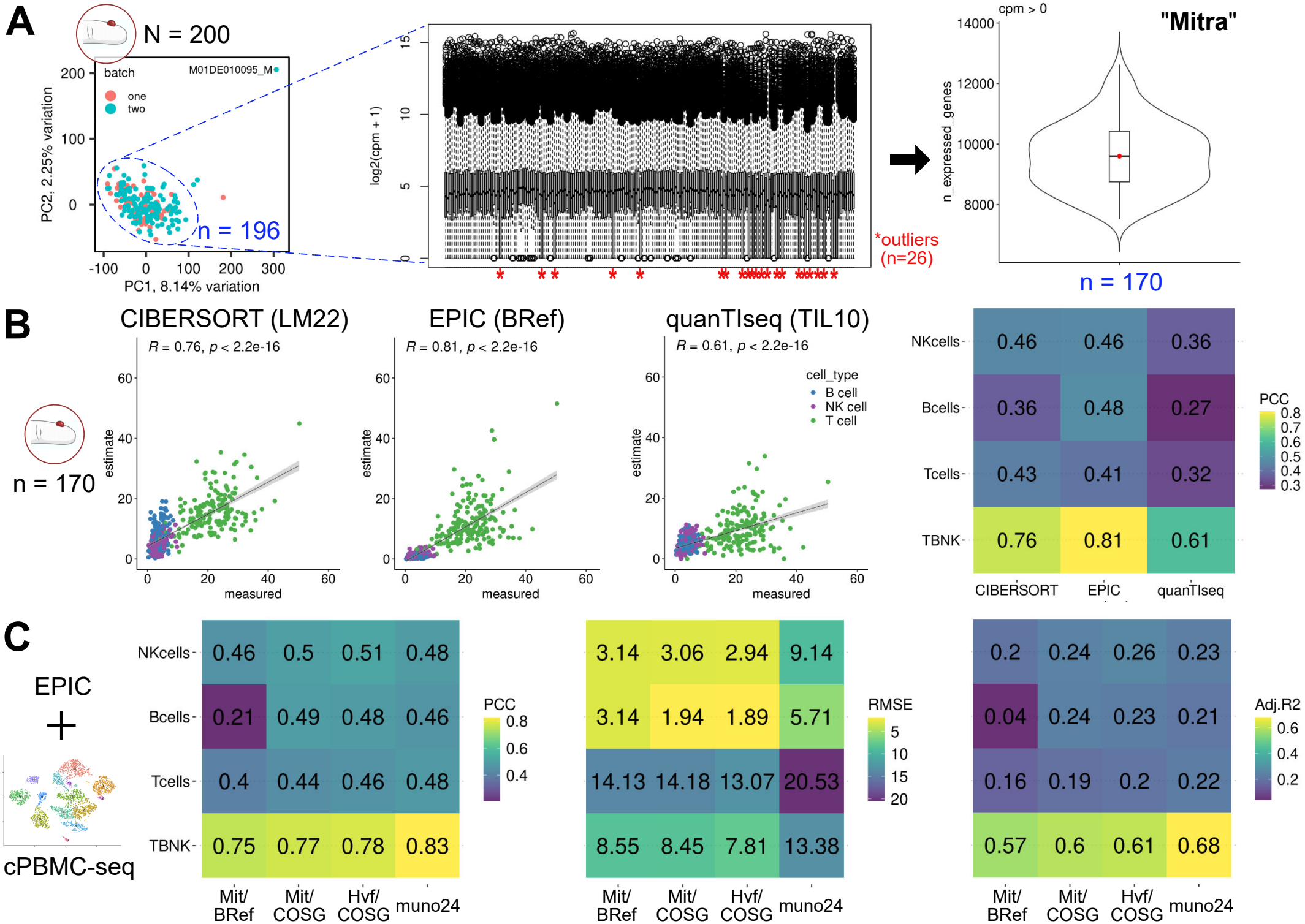
TBNK deconvolution in Mitra dried blood microsamples. **(A)** PCA of dried finger-prick capillary blood (“**Mitra**”) experimental dataset with 4 outliers excluded. Boxplot highlighting asymmetrical skew in the distribution of normalised gene counts (log_2_(CPM + 1)) in an additional 26 outliers. Violin plot of detected genes (CPM *>* 0) in remaining 170 samples, with median (9600 genes) highlighted in red. **(B)** Scatter plots of computed TBNK fractions (“estimate”) vs ground-truth flow cytometry counts (“measured”) using three deconvolution algorithms (CIBERSORT, EPIC, quanTIseq) and their respective pre-packaged signatures (LM22, BRef, TIL10). Pearson’s *r* and *p* value for collective TBNK deconvolution results are labelled. Tile plot summarising performance metrics (PCC; RMSE and Adj.R2 are shown in Supplementary Figure S1A) of TBNK deconvolution split by cell type (“Tcells”, “Bcells”, or “NKcells”) or collectively (“TBNK”), across the three algorithms. **(C)** Tile plots summarising performance metrics of TBNK deconvolution split by cell type (“Tcells”, “Bcells”, or “NKcells”) or collectively (“TBNK”) with EPIC using custom reference profiles computed from a published capillary blood scRNA-seq dataset (“cPBMC-seq”). Mit; cPBMC-seq counts of all genes in Mitra dataset, COSG; 65 TBNK markers computed from cPBMC-seq, Hvf; cPBMC-seq counts of 793 highly variable features, muno24; 24-gene signature identified using our feature selection algorithm, PCC; Pearson Correlation Coefficient, RMSE; Root Mean Squared Error, Adj.R2; Adjusted R-squared, NK; Natural Killer.

Deconvolution with CIBERSORT, EPIC, and quanTIseq showed reduced performance with Mitra-derived RNA compared with PAXgene-derived RNA (Figure 2B). As detailed in the performance heatmap, cell type specific Pearson correlation coefficients did not exceed 0.48 for any method. Correlations calculated across the combined TBNK estimations were higher but differed among the methods (PCC = 0.76 for CIBERSORT, 0.81 for EPIC, and 0.61 for quanTIseq). EPIC provided the most consistent estimates across the three individual cell populations (PCC = 0.41–0.48; RMSE ≤ 12.3 %) (Figure 2B, Supplementary Figure S1A). Nevertheless, its cell type specific performance remained markedly lower than that obtained from the PAXgene dataset.

Applying a stricter filter that retained only the 2592 genes detected in every Mitra sample further reduced EPIC performance (PCC = 0.06–0.37; RMSE ≤ 28.9 %). Conversely, retaining all 25 644 annotated genes did not improve performance relative to the 50 % detection threshold (Supplementary Figure S1B). These findings indicated that the reduced performance in Mitra samples could not be addressed through simple changes to the gene-filtering threshold.

### Capillary-specific reference profiles provide limited improvement in Mitra deconvolution

Previous benchmarking studies have shown that deconvolution performance can be strongly influenced by the choice of reference signature matrix [23, 24]. EPIC’s predefined BRef signature was constructed using gene expression profiles derived from venous peripheral blood. As subtle differences in immune cell composition and transcriptional profiles have been reported between venous and capillary blood [19], we investigated whether a capillary blood specific reference could improve the deconvolution of the Mitra transcriptomes. The EPIC reference framework comprises a gene-by-cell-type expression profile, a corresponding set of cell type marker genes, and experimentally validated estimates of the relative mRNA content of each cell type. We first generated a custom gene-by-cell-type expression profile from a published capillary blood single-cell RNA-seq (scRNA-seq) dataset (cPBMC-seq [19]) and combined it with the original BRef marker gene set. This configuration, termed Mit/BRef, did not improve performance relative to the standard EPIC analysis. B cell prediction was particularly limited (PCC = 0.21; RMSE = 3.1 %; Figure 2C).

We subsequently identified 65 TBNK-specific marker genes from the capillary blood single-cell dataset using COSG [25] (Supplementary Figures S2–S3, Supplementary Table S1). Replacing the BRef markers with these capillary-derived markers (Mit/COSG) improved B cell prediction (PCC = 0.49 versus 0.21; RMSE = 1.9 % versus 3.1 %). However, there was no comparable improvement in T cell or NK cell prediction. Restricting the custom reference profile to 783 highly variable features (Hvf/COSG) produced similar cell type specific correlations (PCC = 0.46–0.51), rather than a substantial additional improvement.

Finally, we evaluated an exploratory 24-gene signature (muno24) identified using a genetic feature-selection procedure (Materials & Methods) applied to the PAXgene dataset. The resulting cell type specific correlations were similar to those obtained using the larger capillary-specific profiles, but prediction error was higher (RMSE ≤ 20.5 %). Although the 24-gene signature produced a strong collective TBNK correlation (PCC = 0.83), it did not consistently improve the accuracy of the individual cell type estimates. Overall, adaptation of the reference profile produced a specific improvement in B cell prediction but did not restore the performance observed with stabilised venous blood.

### Reduced transcriptome complexity contributes to reduced performance in Mitra samples

We next investigated whether the reduced complexity of the Mitra expression profiles contributed to their poorer predictive performance. Stringent gene-retention criteria (raw count > 2 in ≥ 198 of 200 samples) were applied to the raw count matrices of both datasets, retaining 7787 and 1123 genes in the PAXgene and Mitra matrices, respectively. In addition to the lower number of retained genes, Mitra libraries contained fewer total counts per sample than PAXgene libraries (Figure 3A). Linear-regression models trained to predict the flow cytometric fractions, centred at zero within each cell type, showed substantially stronger performance with PAXgene than with Mitra expression data. Across the combined TBNK predictions, the PCC was 0.83 for PAXgene and 0.14 for Mitra (Figure 3A’).

**Figure 3:**
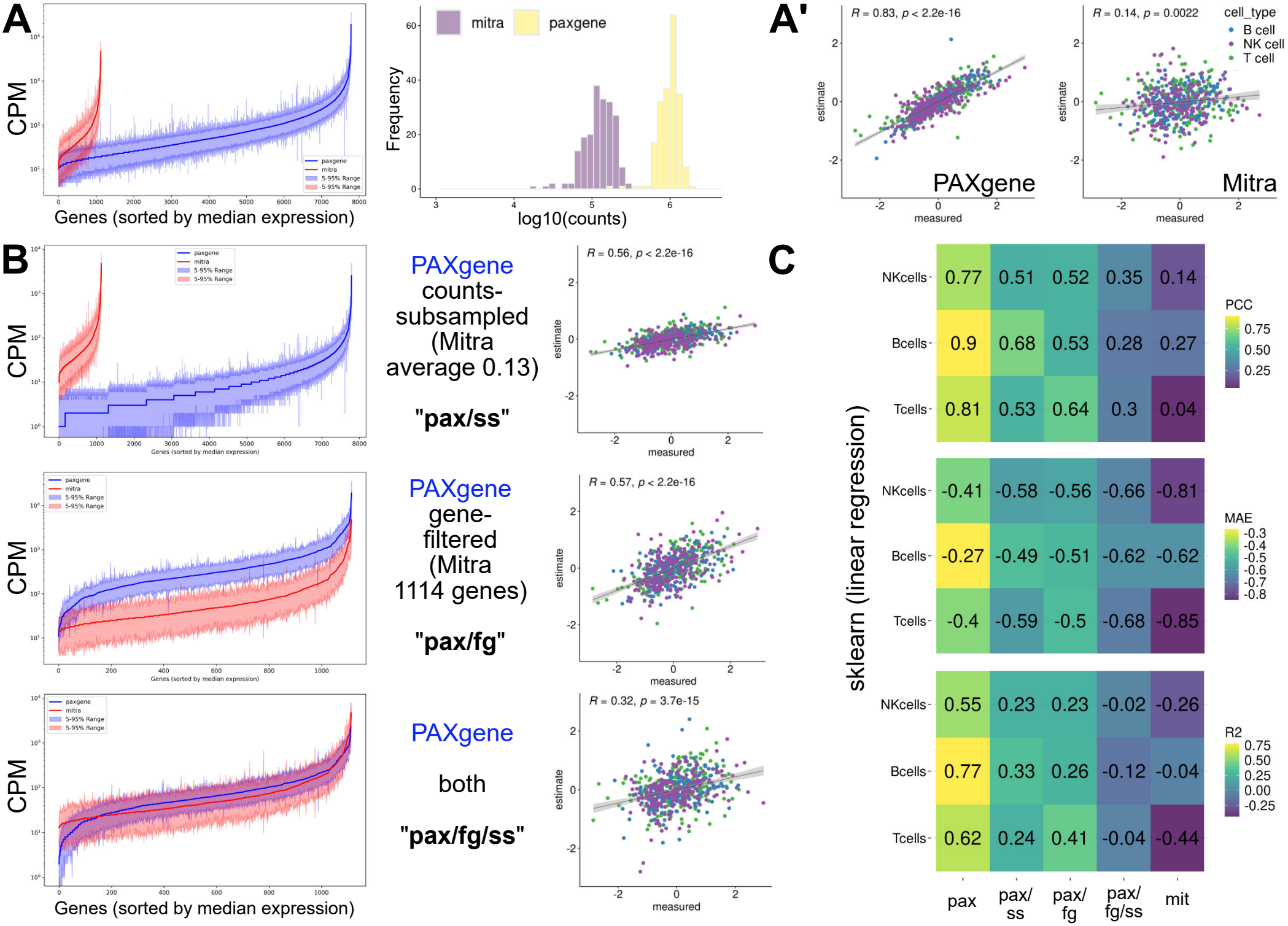
Reduced transcriptome complexity in Mitra samples is associated with poorer TBNK fraction predictions. **(A)** Expression values (CPM) of genes retained after application of stringent gene-retention criteria to the raw PAXgene and Mitra count matrices (raw counts *>* 2 in at least 198 of 200 samples): PAXgene (blue, 7787 genes) and Mitra (red, 1123 genes). Genes are sorted by ascending median expression and shaded areas demarcate the 5–95 % quantiles. Histogram (bins = 50) showing distribution of total counts per sample. **(A’)** Scatter plots of computed TBNK fractions (“estimate”) vs ground-truth flow cytometry fractions (“measured”) using linear regression for the PAXgene andMitra datasets. **(B)** TBNK predictions in PAXgene after counts were subsampled to the mean Mitra/PAXgene total-count ratio (= 0.13), the PAXgene retained genes were restricted to the 1114 genes shared with Mitra, or both modifications were applied. **(C)** Tile plots summarising performance metrics of TBNK predictions split by cell type (“Tcells”, “Bcells”, or “NKcells”). pax; PAXgene retained-gene matrix (7787 genes), ss; counts subsampled to the mean Mitra/PAXgene totalcount ratio (= 0.13), fg; genes restricted to those shared with Mitra (1114 genes), fg/ss; both gene restriction and count subsampling applied, mit; Mitra retained-gene matrix (1123 genes), PCC; Pearson Correlation Coefficient, MAE; Mean Absolute Error, R2; R-squared, NK; Natural Killer.

To model the effect of reduced transcript abundance, PAXgene counts were subsampled to the mean Mitra:PAXgene count ratio of 0.13 (“pax/ss”). This reduced collective TBNK performance (PCC = 0.56 versus 0.83; Figure 3B). Restricting the PAXgene expression matrix to the 1114 genes shared with Mitra produced a similar reduction (PCC = 0.57; Figure 3B) (“pax/fg”). Applying both count subsampling and gene restriction reduced performance further (PCC = 0.32; Figure 3B) (“pax/fg/ss”). Cell type specific correlations were similarly reduced by count subsampling, gene filtering, or both (Figure 3C).

## 1 Discussion

In this study, we investigated the feasibility of using dried capillary blood microsamples for the transcriptomics-based digital cytometry of major circulating immune cells. While our experimental workflow achieved robust deconvolution of TBNK fractions using stabilised venous blood (PAXgene), patient-matched dried capillary blood microsamples (Mitra) exhibited a marked reduction in deconvolution accuracy. Extensive adaptation of the reference profiles, including the integration of capillary-specific single-cell marker genes and custom signatures, provided only limited cell type specific improvements. This indicated that the performance deficit could not be resolved solely through computational adjustments.

Instead, our analyses identified reduced transcriptome complexity as a primary driver of the reduced predictive performance in Mitra samples. Using the same standard library preparation workflow, the dried microsamples yielded libraries with both a lower number of detected genes and a lower average read count per gene compared with stabilised venous samples. By experimentally subsampling read counts and restricting the detectable genes in the PAXgene dataset, we were able to largely recapitulate the deterioration in TBNK fraction predictions. This confirms the critical role of transcript diversity and abundance in successful cellular deconvolution. However, the failure of computational downsampling to fully mirror the severity of the Mitra performance drop suggests that additional biological differences between capillary and venous blood, or other unmeasured pre-analytical variables, may also contribute to the discrepancy.

Several methodological factors inherent to our study design may account for this reduced library complexity. The Mitra devices employ proprietary Volumetric Absorptive Microsampling (VAMS) technology, which immediately dries the post draw blood. While this desiccation is highly beneficial for stabilising messenger RNA at ambient temperatures and facilitating out-of-clinic or home-based sample collection [26, 27, 28], it is possible that alternative microsampling devices might preserve RNA integrity or extraction efficiency more effectively. Furthermore, the diminished signal in the Mitra libraries may be heavily influenced by the low input volume (2 µL) specified by the chosen library preparation protocol.

This low input likely resulted in a dilutive effect on the absolute number and diversity of transcripts captured from the total extracted RNA per sample.

Addressing these technical hurdles will be essential for the future application of digital cytometry to microsampling. Future efforts must focus on generating capillary blood transcriptomes with a complexity on par with those derived from standard venipuncture. Strategies to maximise library complexity could include modifying library preparation protocols to accommodate larger input volumes (e.g., ≥ 10 µL). Additionally, employing full-length RNA-seq approaches, such as SMART-seq [29, 30], may provide a more comprehensive capture of the transcriptome than the 3’-end sequencing approach used here, potentially buffering against the low initial input mass characteristic of microsamples.

The quantification of whole blood immune cell populations remains a routine and critical task for diagnosing and monitoring disease. While flow cytometry is the established gold standard for this purpose, its reliance on fresh tissue, complex logistics, and specialised infrastructure limits its utility in remote and high-frequency longitudinal settings. The convergence of next-generation sequencing, bioinformatic deconvolution, and minimally invasive microsampling offers a powerful alternative. Although our findings highlight that optimising pre-analytical processing and library complexity is a critical prerequisite, overcoming these physical sequencing limitations will ultimately unlock the potential of digital cytometry to democratise and decentralise cellular health monitoring in a digital health era.

## Data Availability

The code used to perform the analyses in this study is available from the
corresponding author upon reasonable request. Individual-level participant
metadata are not publicly available because they contain confidential clinical
information and their disclosure is not permitted under the participant consent
and ethical approval for this study. Aggregate data supporting the findings are
provided within the article and its Supplementary Information.

## Data and code availability

The code used to perform the analyses in this study is available from the corresponding author upon reasonable request. Individual-level participant metadata are not publicly available because they contain confidential clinical information and their disclosure is not permitted under the participant consent and ethical approval for this study. Aggregate data supporting the findings are provided within the article and its Supplementary Information.

## Acknowledgements

We thank the physicians, nurses, and other clinical staff at Medizinicum Hamburg for their support in conducting the study, recruiting participants, and collecting study samples. We are especially grateful to all study participants for their time and contribution to this research.

## Abbreviations

COPD: chronic obstructive pulmonary disease
CPM: counts per million
mRNA: messenger RNA
PCA: principal-component analysis
PCR: polymerase chain reaction
RMSE: root-mean-square error
TBNK: collective term for T cells,B cells,and natural killer cells.

## Author contributions

**Q.R.V.F**.: Conceptualization, Formal analysis, Methodology, Writing – original draft, Writing – review & editing; **T.R**.: Conceptualization, Investigation, Methodology, Writing – original draft, Writing – review & editing; **M.Z**.: Investigation, Project administration, Resources, Writing – review & editing; **V.C.M**.: Formal analysis, Investigation, Visualization, Writing – original draft, Writing – review & editing; **T.v.S**.: Resources, Supervision, Writing – review & editing; **K.A.-S**.: Investigation, Project administration, Resources, Supervision, Writing – review & editing; **B.R.S**.: Conceptualization, Investigation, Methodology, Project administration, Resources, Supervision, Funding acquisition, Writing – original draft, Writing – review & editing;

## Supplementary Figures

**Supplementary Figure S1:**
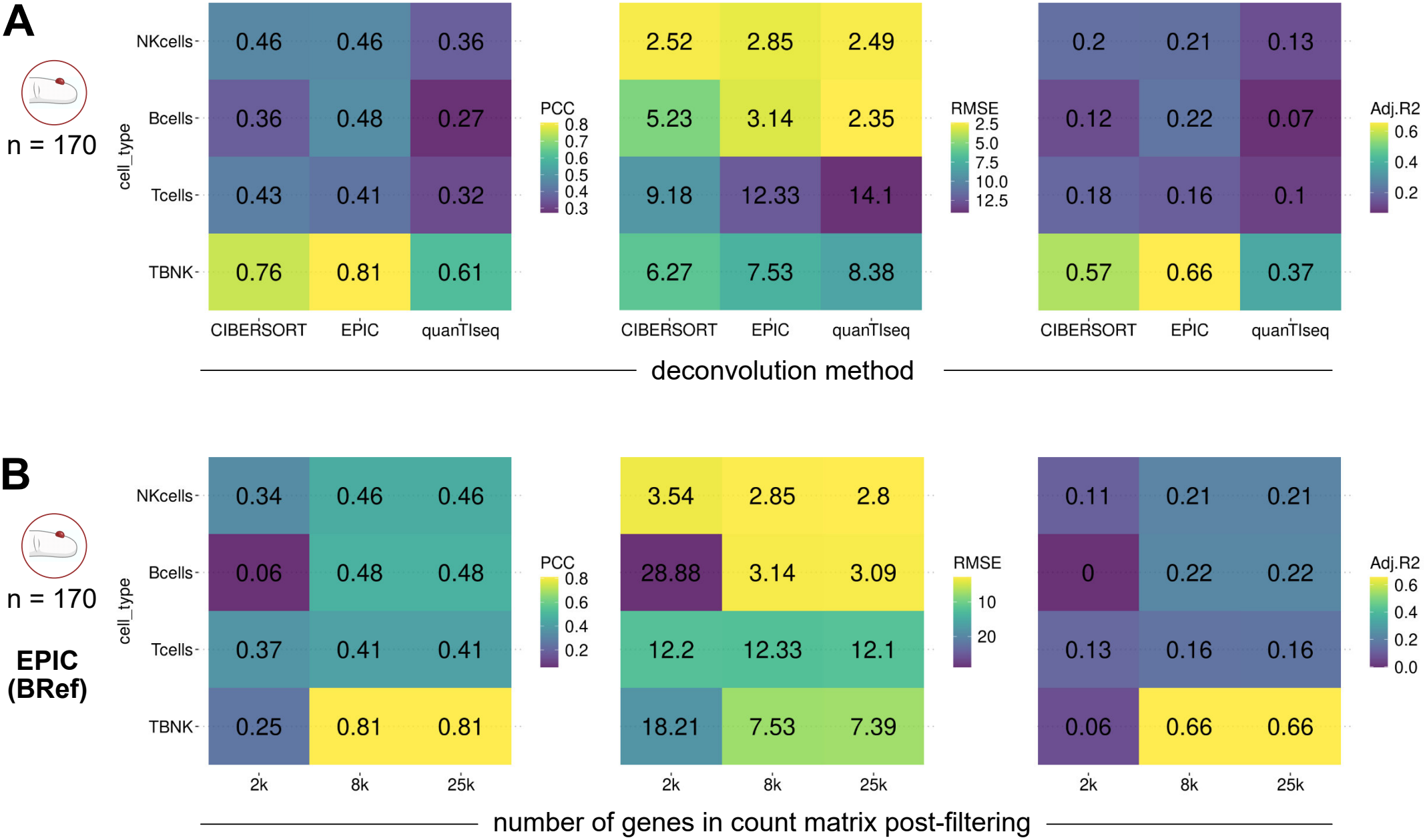
Mitra TBNK deconvolution. **(A)** Tile plot summarising performance metrics of Mitra TBNK deconvolution split by cell type (“Tcells”, “Bcells”, or “NKcells”) or collectively (“TBNK”) by CIBERSORT, EPIC, and quanTIseq. **(B)** Tile plot summarising performance metrics of TBNK deconvolution (EPIC) with three gene-filtering thresholds applied to Mitra.

**Supplementary Figure S2:**
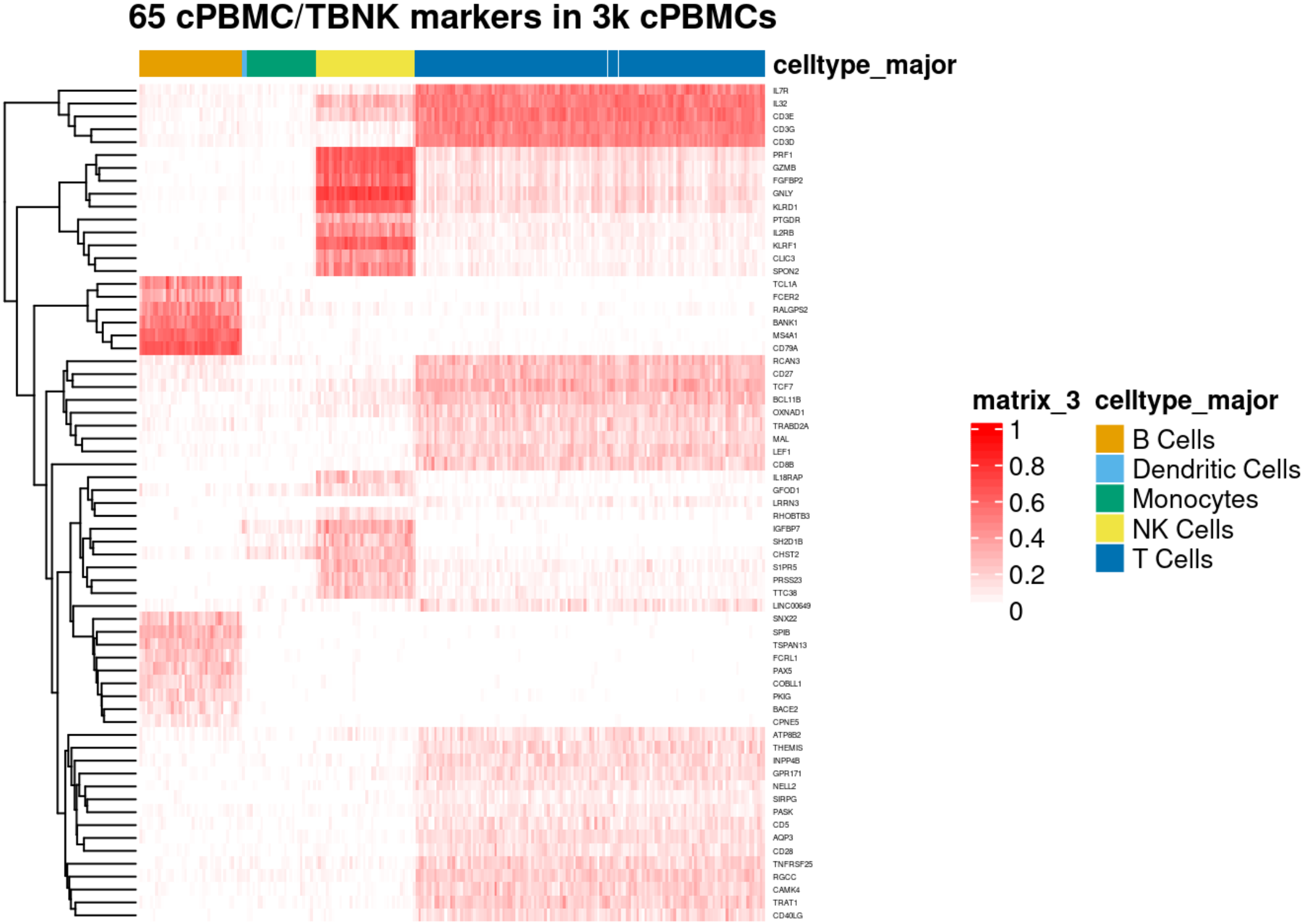
Expression of COSG.TBNK.Mitra marker genes in capillary blood scRNA-seq. 65 TBNK-only marker genes from capillary blood scRNA-seq were identified using COSG [25].

**Supplementary Figure S3:**
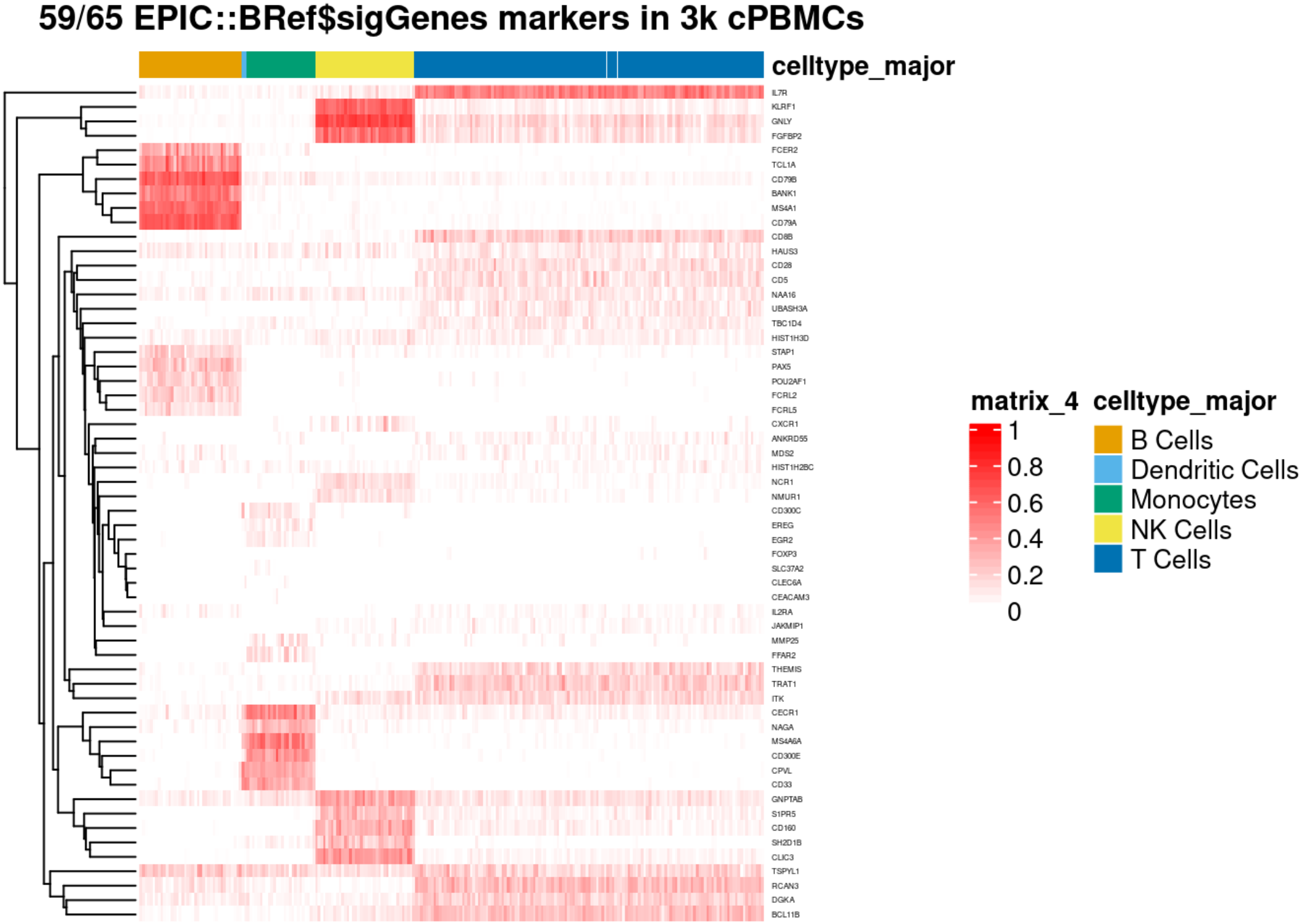
Expression of EPIC::BRef$sigGenes marker genes in capillary blood scRNA-seq. 59 out of the total 65 marker genes were detected in capillary blood scRNA-seq.

## Supplementary Tables

**Supplementary Table S1:** TBNK gene markers for EPIC deconvolution. `COSG.TBNK.Mitra` was derived from capillary blood and `EPIC/BRef$sigGenes` from peripheral blood.

|  | COSG.TBNK.Mitra |  | EPIC::BRef\$sigGenes<sup>*</sup> |
|---|---|---|---|
| **T cells<sup>†</sup>** | **CD5** | OXNAD1 | **BCL11B** |
|  | CD3G | RGCC | **CD5** |
|  | CD3D | SIRPG | **CD28** |
|  | TRAT1 | CD27 | **IL7R** |
|  | CD40LG | GPR171 | ITK |
|  | **THEMIS** | TNFRSF25 | **THEMIS** |
|  | **IL7R** | LRRN3 | UBASH3A |
|  | CD8B | TCF7 | – |
|  | MAL | CD3E | – |
|  | CAMK4 | TRABD2A | – |
|  | LEF1 | LINC00649 | – |
|  | AQP3 | **BCL11B** | – |
|  | INPP4B | RCAN3 | – |
|  | **CD28** | IL32 | – |
|  | NELL2 | ATP8B2 | – |
|  | PASK | – | – |
| **B cells<sup>†</sup>** | **MS4A1** | PKIG | **BANK1** |
|  | **CD79A** | RALGPS2 | **CD79A** |
|  | **BANK1** | BACE2 | CD79B |
|  | **TCL1A** | CPNE5 | **FCER2** |
|  | SPIB | – | FCRL2 |
|  | **FCER2** | – | FCRL5 |
|  | TSPAN13 | – | **MS4A1** |
|  | **PAX5** | – | **PAX5** |
|  | FCRL1 | – | POU2AF1 |
|  | COBLL1 | – | STAP1 |
|  | SNX22 | – | **TCL1A** |
| **NK cells<sup>†</sup>** | **SH2D1B** | **KLRF1** | CD160 |
|  | **CLIC3** | IL18RAP | **CLIC3** |
|  | SPON2 | IGFBP7 | **FGFBP2** |
|  | RHOBTB3 | IL2RB | **GNLY** |
|  | GZMB | PTGDR | GNPTAB |
|  | PRSS23 | CHST2 | **KLRF1** |
|  | TTC38 | GFOD1 | NCR1 |
|  | **GNLY** | **FGFBP2** | NMUR1 |
|  | **S1PR5** | KLRD1 | **S1PR5** |
|  | PRF1 | – | **SH2D1B** |

<sup>\*</sup> 28 TBNK markers; full list from Racle *et al.* 2017 (Appendix 1 – table 1) consists of 65 markers across 10 cell types including TBNK and others
<sup>†</sup> Common markers between the two gene marker sets are highlighted in **bold**.

## References

[1] Francisco Avila Cobos, Jo Vandesompele, Pieter Mestdagh, and Katleen De Preter. Computational deconvolution of transcriptomics data from mixed cell populations. Bioinformatics, 34(11):1969–1979, 6 2018.

[2] Kobe De Ridder, Huiwen Che, Kaat Leroy, and Bernard Thienpont. Benchmarking of methods for DNA methylome deconvolution. Nature Communications, 15(1):4134, 5 2024.

[3] Aaron M Newman, Chih Long Liu, Michael R Green, Andrew J Gentles, Weiguo Feng, Yue Xu, Chuong D Hoang, Maximilian Diehn, and Ash A Alizadeh. Robust enumeration of cell subsets from tissue expression profiles. Nature Methods, 12(5):453–457, 5 2015.

[4] Julien Racle, Kaat de Jonge, Petra Baumgaertner, Daniel E Speiser, and David Gfeller. Simultaneous enumeration of cancer and immune cell types from bulk tumor gene expression data. eLife, 6, 11 2017.

[5] Francesca Finotello, Clemens Mayer, Christina Plattner, Gerhard Laschober, Dietmar Rieder, Hubert Hackl, Anne Krogsdam, Zuzana Loncova, Wilfried Posch, Doris Wilflingseder, Sieghart Sopper, Marieke Ijsselsteijn, Thomas P. Brouwer, Douglas Johnson, Yaomin Xu, Yu Wang, Melinda E. Sanders, Monica V. Estrada, Paula Ericsson-Gonzalez, Pornpimol Charoentong, Justin Balko, Noel Filipe da Cunha Carvalho de Miranda, and Zlatko Trajanoski. Molecular and pharmacological modulators of the tumor immune contexture revealed by deconvolution of RNA-seq data. Genome Medicine, 11(1):34, 12 2019.

[6] Gianni Monaco, Bernett Lee, Weili Xu, Seri Mustafah, You Yi Hwang, Christophe Carré, Nicolas Burdin, Lucian Visan, Michele Ceccarelli, Michael Poidinger, Alfred Zippelius, João Pedro de Magalhães, and Anis Larbi. RNA-Seq Signatures Normalized by mRNA Abundance Allow Absolute Deconvolution of Human Immune Cell Types. Cell Reports, 26(6):1627–1640, 2 2019.

[7] Gregory J Hunt, Saskia Freytag, Melanie Bahlo, and Johann A Gagnon-Bartsch. dtangle: accurate and robust cell type deconvolution. Bioinformatics, 35(12):2093–2099, 6 2019.

[8] Elizabeth H Robison, Tony S Mondala, Adam R Williams, Steven R Head, Daniel R Salomon, and Sunil M Kurian. Whole genome transcript profiling from fingerstick blood samples: a comparison and feasibility study. BMC Genomics, 10(1):617, 12 2009.

[9] Timothy M. Blicharz, Ping Gong, Bernard M. Bunner, Larry L. Chu, Kaela M. Leonard, Jessica A. Wakefield, Richard E. Williams, Maisam Dadgar, Carlo A. Tagliabue, Ragheb El Khaja, Stephanie L. Marlin, Ramin Haghgooie, Shawn P. Davis, Donald E. Chickering, and Howard Bernstein. Microneedle-based device for the one-step painless collection of capillary blood samples. Nature Biomedical Engineering, 2(3):151–157, 2 2018.

[10] Alexis Catala, Rachel Culp-Hill, Travis Nemkov, and Angelo D’Alessandro. Quantitative metabolomics comparison of traditional blood draws and TAP capillary blood collection. Metabolomics, 14(7):100, 7 2018.

[11] Joost Brandsma, Josh G. Chenoweth, Melissa K. Gregory, Subramaniam Krishnan, Paul W. Blair, Deborah A. Striegel, Rittal Mehta, Kevin L. Schully, J. Stephen Dumler, CDR Cynthia S. Sikorski, Kelsey O’Connor, Susan A. Reichert-Scrivner, Carmen M. Paguirigan, Catherine F. T. Uyehara, COL Viseth Ngauy, Christopher A. Myers, and Danielle V. Clark. Assessing the use of a micro-sampling device for measuring blood protein levels in healthy subjects and COVID-19 patients. PLOS ONE, 17(8):e0272572, 8 2022.

[12] Amanda J. Campbell, Nicolai B. Palstrøm, Lars M. Rasmussen, Jes S. Lindholt, and Hans C. Beck. From blood drops to biomarkers: a scoping review of microsampling in mass spectrometry-based proteomics. Clinical Proteomics, 22(1):20, 12 2025.

[13] Abbie D. Leino, John Takyi-Williams, Jeong M. Park, Silas P. Norman, Duxin Sun, Karen B. Farris, and Manjunath P. Pai. Clinical validation of two volumetric absorptive microsampling devices to support home-based therapeutic drug monitoring of immunosuppression. British Journal of Clinical Pharmacology, 90(11):2897–2909, 11 2024.

[14] Arkadiusz Kocur and Tomasz Pawiński. Microsampling techniques and patient-centric therapeutic drug monitoring of immunosuppressants. Bioanalysis, 17(6):413–427, 3 2025.

[15] Ryan Toma, Nathan Duval, Benjamin Pelle, Matthew M Parks, Vishakh Gopu, Pedro J Torres, Francine R Camacho, Nan Shen, Subha Krishnan, Andrew Hatch, Hal Tily, Ally Perlina, Guruduth Banavar, and Momchilo Vuyisich. A Clinically Validated Human Capillary Blood Transcriptome Test For Global Systems Biology Studies. BioTechniques, 69(4):289–301, 10 2020.

[16] Benjamin Kaminow, Dinar Yunusov, and Alexander Dobin. STARsolo: accurate, fast and versatile mapping/quantification of single-cell and single-nucleus RNA-seq data. bioRxiv, 5 2021.

[17] Mark D Robinson and Alicia Oshlack. A scaling normalization method for differential expression analysis of RNA-seq data. Genome Biology, 11(3):R25, 3 2010.

[18] Gregor Sturm, Francesca Finotello, Florent Petitprez, Jitao David Zhang, Jan Baumbach, Wolf H Fridman, Markus List, and Tatsiana Aneichyk. Comprehensive evaluation of transcriptome-based cell-type quantification methods for immuno-oncology. Bioinformatics, 35(14):i436–i445, 7 2019.

[19] Tatyana Dobreva, David Brown, Jong Hwee Park, and Matt Thomson. Single cell profiling of capillary blood enables out of clinic human immunity studies. Scientific Reports, 10(1):20540, 11 2020.

[20] Yuhan Hao, Tim Stuart, Madeline H. Kowalski, Saket Choudhary, Paul Hoffman, Austin Hartman, Avi Srivastava, Gesmira Molla, Shaista Madad, Carlos Fernandez-Granda, and Rahul Satija. Dictionary learning for integrative, multimodal and scalable single-cell analysis. Nature Biotechnology, 42(2):293–304, 2 2024.

[21] Christoph Hafemeister and Rahul Satija. Normalization and variance stabilization of single-cell RNA-seq data using regularized negative binomial regression. Genome Biology, 20(1):296, 12 2019.

[22] Fabian Pedregosa, Gael Varoquaux, Alexandre Gramfort, Vincent Michel, Bertrand Thirion, Olivier Grisel, Mathieu Blondel, Peter Prettenhofer, Ron Weiss, Vincent Dubourg, Jake Vanderplas, Alexandre Passos, David Cournapeau, Matthieu Brucher, Matthieu Perrot, and Édouard Duchesnay. Scikit-learn: Machine learning in Python. Journal of Machine Learning Research, 12, 2011.

[23] Francisco Avila Cobos, José Alquicira-Hernandez, Joseph E. Powell, Pieter Mestdagh, and Katleen De Preter. Benchmarking of cell type deconvolution pipelines for transcriptomics data. Nature Communications, 11(1):5650, 11 2020.

[24] Brian B Nadel, Meritxell Oliva, Benjamin L Shou, Keith Mitchell, Feiyang Ma, Dennis J Montoya, Alice Mouton, Sarah Kim-Hellmuth, Barbara E Stranger, Matteo Pellegrini, and Serghei Mangul. Systematic evaluation of transcriptomics-based deconvolution methods and references using thousands of clinical samples. Briefings in Bioinformatics, 22(6), 11 2021.

[25] Min Dai, Xiaobing Pei, and Xiu-Jie Wang. Accurate and fast cell marker gene identification with COSG. Briefings in Bioinformatics, 23(2), 3 2022.

[26] Yoshimasa Maeno, Shusuke Nakazawa, Shigeo Nagashima, Jun Sasaki, Kyoko M. Higo, and Koki Taniguchi. Utility of the dried blood on filter paper as a source of cytokine mRNA for the analysis of immunoreactions in Plasmodium yoelii infection. Acta Tropica, 87(2):295–300, 7 2003.

[27] Fredrika Gauffin, Ann Nordgren, Gisela Barbany, Britt Gustafsson, and Håkan Karlsson. Quantitation of RNA decay in dried blood spots during 20 years of storage. Clinical Chemistry and Laboratory Medicine, 47(12), 1 2009.

[28] Mary J. Reust, Myung Hee Lee, Jenny Xiang, Wei Zhang, Dong Xu, Tatiana Batson, Tuo Zhang, Jennifer A. Downs, and Kathryn M. Dupnik. Dried Blood Spot RNA Transcriptomes Correlate with Transcriptomes Derived from Whole Blood RNA. The American Journal of Tropical Medicine and Hygiene, 98(5):1541–1546, 5 2018.

[29] Daniel Ramsköld, Shujun Luo, Yu-Chieh Wang, Robin Li, Qiaolin Deng, Omid R Faridani, Gregory A Daniels, Irina Khrebtukova, Jeanne F Loring, Louise C Laurent, Gary P Schroth, and Rickard Sandberg. Full-length mRNA-Seq from single-cell levels of RNA and individual circulating tumor cells. Nature Biotechnology, 30(8):777–782, 8 2012.

[30] Simone Picelli, Åsa K Björklund, Omid R Faridani, Sven Sagasser, Gösta Winberg, and Rickard Sandberg. Smart-seq2 for sensitive full-length transcriptome profiling in single cells. Nature Methods, 10(11):1096–1098, 11 2013.

